# Modeling COVID-19 epidemics in an Excel spreadsheet: Democratizing the access to first-hand accurate predictions of epidemic outbreaks

**DOI:** 10.1101/2020.03.23.20041590

**Authors:** Mario Moisés Alvarez, Everardo González-González, Grissel Trujillo-de Santiago

**Affiliations:** Centro de Biotecnología-FEMSA, Tecnologico de Monterrey, Monterrey 64849, NL, México; Departamento de Bioingeniería, Escuela de Ingeniería y Ciencias, Tecnologico de Monterrey, Monterrey 64849, NL, México; Departamento de Ingeniería Mecatrónica y Eléctrica, Escuela de Ingeniería y Ciencias, Tecnologico de Monterrey, Monterrey 64849, NL, México

**Keywords:** COVID-19, coronavirus, SARS-CoV2, mathematical modeling, epidemic, pandemic, Excel

## Abstract

COVID-19, the first pandemic of this decade and the second in less than 15 years, has harshly taught us that viral diseases do not recognize boundaries; however, they truly do discriminate between aggressive and mediocre containment responses.

We present a simple epidemiological model that is amenable to implementation in Excel spreadsheets and sufficiently accurate to reproduce observed data on the evolution of the COVID-19 pandemics in different regions (i.e., Italy, Spain, and New York City (NYC)). We also show that the model can be adapted to closely follow the evolution of COVID-19 in any large city by simply adjusting two parameters related to (a) population density and (b) aggressiveness of the response from a society/government to epidemics. Moreover, we show that this simple epidemiological simulator can be used to assess the efficacy of the response of a government/society to an outbreak.

The simplicity and accuracy of this model will greatly contribute to democratizing the availability of knowledge in societies regarding the extent of an epidemic event and the efficacy of a governmental response.

## Introduction

A SARS-CoV2 (COVID-19) pandemic was declared by the World Health Organization in March 2020. More than 100,000 positive cases of COVID-19 infection had been declared worldwide at that point, mainly in China, Italy, Iran, Spain, and other European countries. By the end of March 2020, the official cumulative number of infected worldwide ascended to more than 700,000, with a toll of death higher than 32,000 and a strong presence in Las Americas, mainly in the USA^1^. COVID-19, the first pandemic of this decade and the second in less than 15 years, has harshly taught us that viral diseases do not recognize boundaries; however, they truly do discriminate between aggressive and mediocre containment responses.

Indeed, three months have passed since the emergence of COVID-19, and we have been able to observe exemplary responses from some Asian countries (i.e., China^2^, South Korea^3^, Singapore^4^, and Japan), some highly aggressive responses in Europe and America (i.e., Germany and USA), and several delayed or not so effective responses from other regions (i.e., Italy and Spain)^5^. At this point, some territories in Latin America are just experiencing the “lag phase” of the COVID-19 pandemic at home and do not appear having yet implemented proper containment measures as rapidly as needed.

The gap between developed and developing countries may explain some of the differences in the scale of the responses that we are observing. Countries that are better equipped than others in terms of high-end scientific development, diagnostics technology, and health care infrastructure may respond more efficaciously to a pandemic scenario. However, other tools, such as mathematical modeling, are much more widely available and may be of extraordinary value when managing epidemic events such as the COVID-19 pandemics. To date, many papers have reported the use of mathematical models and simulators to evaluate the progression of COVID-19 in local or more global settings^63,7–9^. Predictions on the possible evolution of COVID-19 based on mathematical modeling could therefore represent important tools for designing and/or evaluating countermeasures^8,10–12^.

However, mathematical modeling may (and probably should) become a much more available tool in the case of public health emergencies—one ideally widely available to practically any citizen in any of our societies. One decade ago, during the influenza pandemics, mathematical modeling of epidemic events was the realm of privileged epidemiologists who had (a) a fast computer, (b) programing experience, and (c) and access to epidemiological data. Today, those three ingredients are now reduced to a convectional laptop, very basic differential equation-solving skills, and access to a website with reliable online statistical information on the epidemics.

The main purpose of this contribution is to demonstrate that a simple mathematical model, amenable to implementation in an Excel spreadsheet, can accurately predict the evolution of an epidemic event at a local level (i.e., in any major urban area). This may be extremely valuable for government officials who must predict, with high fidelity, the progression of an epidemic event to better design their action strategies. Moreover, the democratization of the modeling of complex epidemic events will empower citizens, enabling them to forecast, decide, and evaluate. For instance, using this simple model, virtually any citizen could assess, in real time, the efficacy of the actions of her/his society in the face of an outbreak.

## Rationale of the model formulation

Here, we construct a very simple epidemiological model for the propagation of COVID-19 in urban areas. The model is based on a set of differential equations. The first equation of the set (equation 1) states that the rate of accumulation of infected habitants in an urban area (assumed to be a closed system) is given by the sum of the number of new infections (positive contribution), the number of recovered patients (negative contribution), and the number of deaths (negative contribution). A second differential equation states that the rate of accumulation of the infected but asymptotic population is proportional to the population of infected and symptomatic subjects (equation 2). Two additional equations relate the number of deaths and recovered patients with the number of newly infected ones (equation 3 and 4). Finally, the rate of depletion of the pool of the population susceptible to infection is given by the sum of recovered patients, asymptomatic infected, and deaths (equation 5). Recent experimental evidence suggests that rhesus macaques that recovered from SARS-CoV-2 infection could not be reinfected^13^. However, at this point, the acquisition of full immunity to reinfection has not been proved in humans, although it is well documented for other coronavirus infections, such as SARS, and MERS^14,15^. The analysis of sera of one COVID-19 patient showed a peak production of specific IgGs against SARS-COV-2 by two weeks after the onset of symptoms ^16^. Based on immunological information on SARS and MERS epidemiology and the limited evidence on the nature of the host immune response to SARS-COV-2, we assume here that recovered patients become immune to reinfection.

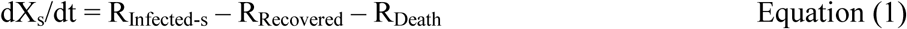

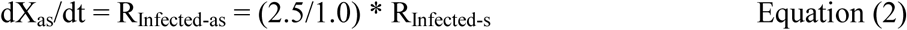

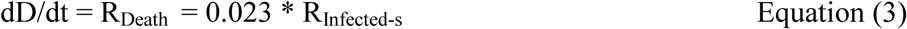

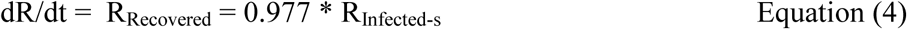

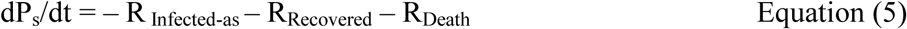

This system is equivalent to

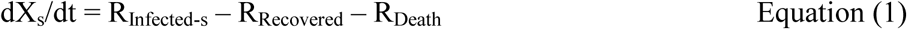

where

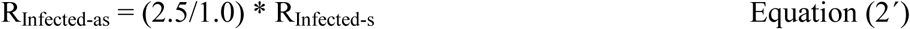

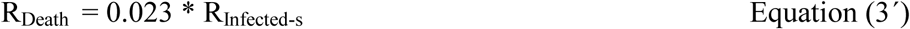

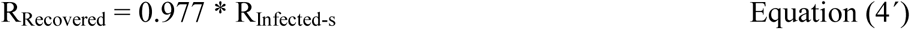

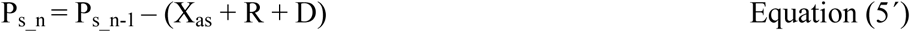

In this system, all equations depend on R_Infected-s._ Here, we propose a simple formulation for the evaluation of R_Infected-s_ at the onset of a local epidemic event.

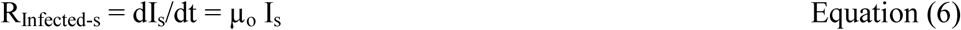

where µ_o_ is the specific rate of infection of a population in a large and vastly uninfected urban area. We further propose that µ_o_ may be calculated from actual epidemiological data corresponding to the first exponential stage of COVID-19 local epidemics. We determined the appropriate ranges of values for µ_o_ by analyzing publicly available data from different websites that continuously monitor the progression of confirmed cases of COVID-19 for different nations (Table 1).

**Table 1.**
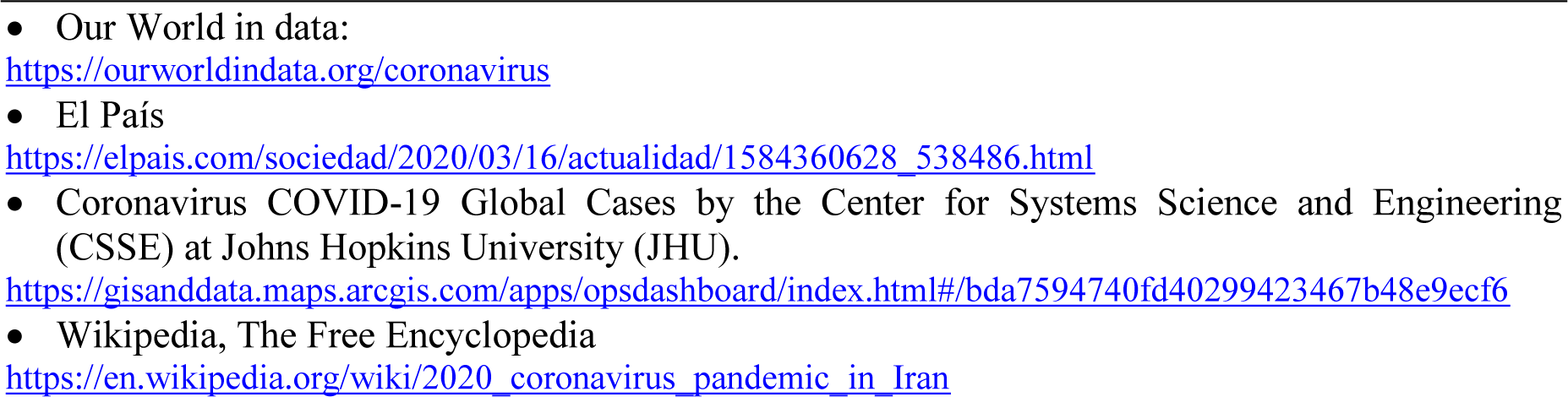
Websites displaying COVID-19 data in practically real time.

This model correctly describes the evolution of the number of newly infected during the initial stage of the epidemic episode. For later times, the rate of new infections is corrected by a term that depends on the demographic density (Dd) of the region. Therefore:

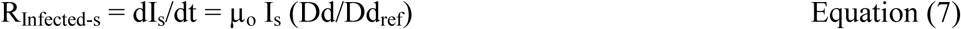

In equation (7), Dd=P_s_/A, where A is the surface area of the region subject to analysis. In this formulation, Dd is the total number of inhabitants of the region who are susceptible to infection, while Dd_ref_ is a value of demographic density in a densely populated urban area that the model uses as a reference. In this work, the demographic density of the city of Madrid is used as Dd_ref_. Furthermore, since Dd is a function that considers only the population susceptible to infection, a counter is needed to continuously update the number of recovered patients, asymptomatic patients, and deaths. Therefore, at each time step during the numerical integration, the susceptible population is updated by subtracting the number of number of recovered patients, asymptomatic patients, and deaths. Note that in our Excel spreadsheet, we use Dd/Dd_ref_ = density_Factor (Supplemental Excel File 1).

Defining an expression for R_Infected-s_ enables stepwise numerical integration, for example by the Euler method. We have implemented this solution in a spreadsheet. To that aim, differential equations (1) and (7) should be converted into their corresponding equations of differences:

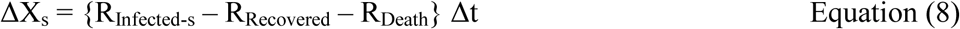

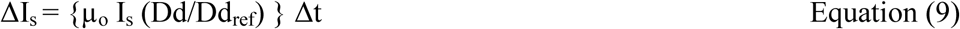

For all the simulation results presented here, we set Δt=1h= 1/24 day. We have solved this differential set, step by step, updating the values of R_Infected-s_, R_Recovered_, R_Death_, and P_s_, according to equations (2’) to (5’). The ratio (Dd/Dd_ref_) is also recalculated at each time step using the updated value of P_s_ from equation (5’).

### Selection of relevant epidemiological parameters for COVID-19

The number of asymptomatic inhabitants was calculated under the assumption that only ∼30.0% of the infected population develops symptomatology (2.5 asymptomatic subjects per 1.0 symptomatic subject). This assumption should be regarded as speculative, since very limited information specific for the ration between symptomatic and asymptomatic COVID-19 patients is available at this point.^17,18^ The percentage of asymptomatic infections during pandemic Influenza A/H1N1/2009, based on epidemiology studies founded in serological analysis in a vast range of geographical settings, has been estimated has been between 65 and 85%^19^. These values are also consistent with the high number of asymptomatic infected subjects estimated for other pandemic events. For instance, in the context of pandemic influenza A/H1N1/2009, up to 20–40% of the population in urban areas (i.e., Monterrey, México, and Pittsburgh, USA) ^20,21^ exhibited specific antibodies regardless of experiencing symptoms, while the fraction of confirmed symptomatic infections was lower than less than 10%.

In addition, the average time of sickness was set at 14 days in our simulations, within the range reported from 14 to 32 days^22^, with a median time to recovery of 21 days^23^. Therefore, the number of patients recovered (R) is calculated as a fraction of 0.977 of those infected 14 days previously. Similarly, asymptomatic patients are only removed from the pool of susceptible after full recovery. Note that, in the current version of our model, asymptomatic patients are not considered part of the population capable of transmitting COVID-19, despite recently reported evidence that suggests that asymptomatic subjects (or minimally symptomatic patients) may exhibit similar viral loads^24^ to those of symptomatic patients and may be active transmitters of the disease^2,25^. The number of deceased patients was calculated as 0.023 of those infected 14 days before. This mortality percentage (case fatality rate) lies within the range reported in recent literature for COVID-19^9,26–28^. The time lapse of 14 days between the onset of disease and death was statistically estimated by Linton et al. in a recent report ^29^.

The straightforward implementation of the model in Excel (Supplemental Excel File 1), using the set of parameters described before, allows the calculation of all populations (I_s,_ X_s_, X_as_, D, R, and P_s_) every hour. Note that this model enables the description of the progressive exhaustion of the epidemic, as expected by the progressive depletion of the susceptible population. Next, we discuss criteria for selection of the values of µ_o_ based on the initial behavior of the COVID-19 Pandemic at different urban areas around the globe.

### Estimation of specific epidemic rate values

Figure 1A shows the progression on the number of COVID-19 positive cases in different regions, namely Spain (mainly Madrid), Iran (mainly Tehran), and New York City (NYC). We have selected these three data sets to illustrate that the evolution of the epidemic has a local flavor that mainly depends on the number of initial infected persons, the demographic density, and the set of containment measures taken by government officials and society. Figure 1B shows the natural log of the cumulative number of infections over time for the same set of countries. This simple plotting strategy is highly useful for analyzing the local rate of progression of the pandemic. If the local epidemic progression is consistent with a simple first order exponential model where dI/dt = µ*I, then the integral form of this equation renders the linear equation: ln I/I_o_= µ*t. During the exponential phase, a straight line should be observed, and the slope of that line denotes the specific rate (µ) of the epidemic spreading. Note that COVID-19 has exhibited a wide range of spreading rates in different countries (from ∼0.3 to ∼0.9 day^-1^). Note also that µ is related to the doubling time (t_d_), often reported in population and epidemiological studies, by the equation t_d_=Ln 2/ µ.

**Figure 1.**
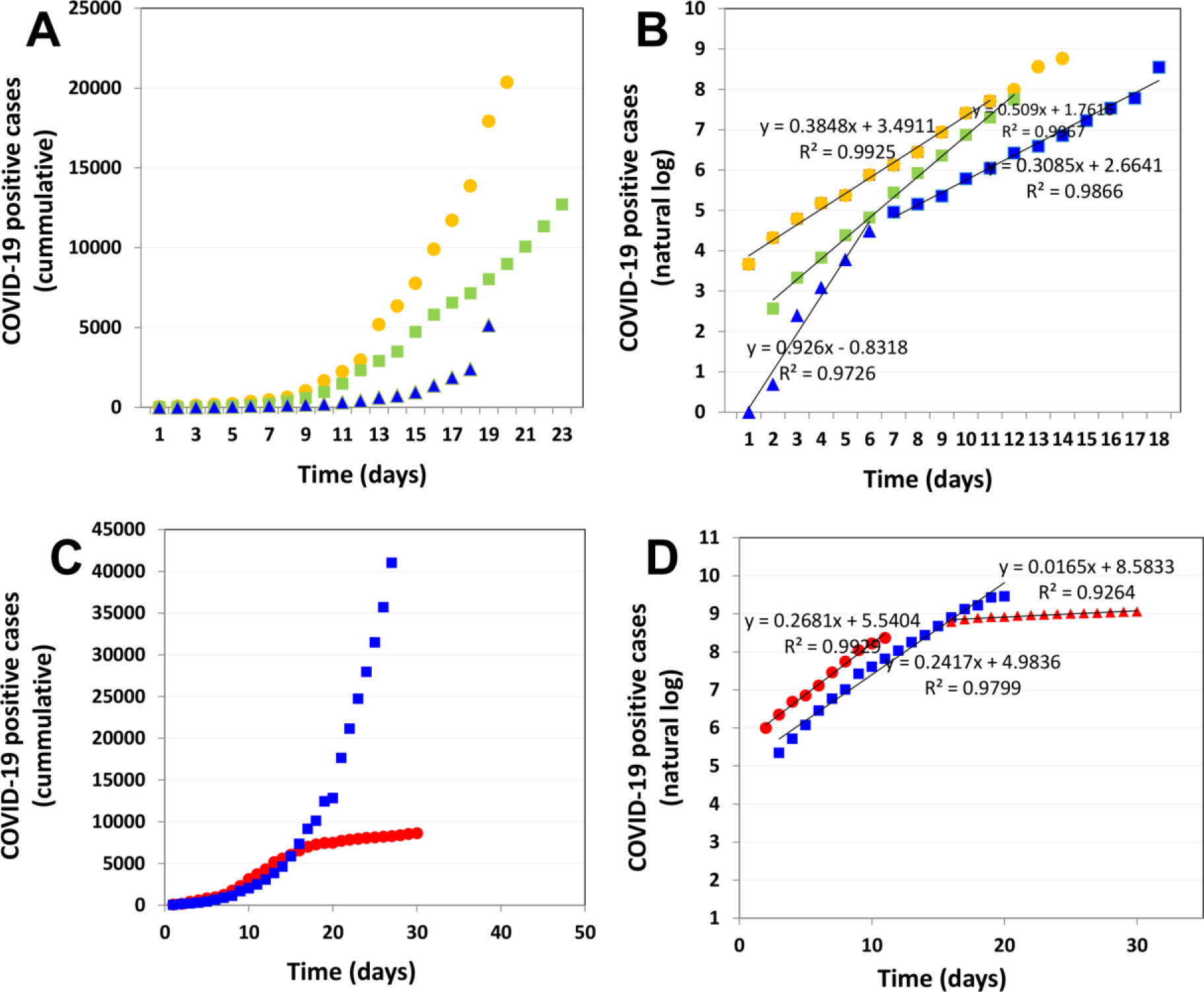
Epidemiological data related to the onset of a COVID-19 pandemic in different regions. (A) Cumulative number of positive cases of COVID-19 infection in Spain (yellow circles), Iran (green squares), and NYC (blue triangles) during the first days after the outbreak. (B) Natural logarithm of the cumulative number of positive cases of COVID-19 infection in Spain (yellow circles), Iran (green squares), and NYC (blue triangles). (C) Cumulative number of positive cases of COVID-19 infection in Italy (blue squares) and South Korea (red circles). (D) Natural logarithm of the cumulative number of positive cases of COVID-19 infection in Italy (blue squares) and South Korea (red circles). Two clearly distinctive exponential stages are observed in the case of South Korean progression.

Therefore, ranges of doubling times between 0.75 and 2.45 days are observed just among these three regional cases.

Different exponential stages, perfectly distinguishable by their exhibition of different slopes (Table 2), may be observed within the same time series. For instance, the outbreak in NYC (Figure 1B; blue symbols) was first described by an extremely high slope (µ_o_ = 0.926 day^-1^). However, after a series of measures adopted in NYC by the federal, state, and local governments, the specific growth rate of the epidemics fell to µ = 0.308 day^-1^.

**Table 2.** Specific infection rates (µ_o_) and associated doubling times (t_d_) for COVID-19 infection in different geographic regions.

| Territory | Temporality | $\mu$ | $t_d$ |
| --- | --- | --- | --- |
| Spain (Madrid) | initial | 0.358 | 1.937 |
| Italy | initial | 0.326 | 2.128 |
| Italy | after stringent measures | 0.119 | 5.849 |
| Iran | initial | 0.491 | 1.411 |
| Iran (Tehran) | initial | 0.506 | 1.370 |
| Germany | initial | 0.280 | 2.474 |
| NYC | initial | 0.591 | 1.173 |
| NYC | after measures | 0.293 | 2.362 |
| South Korea | initial | 0.293 | 2.362 |
| South Korea | after stringent measures; massive testing | 0.000 | ND* |
| France | initial | 0.379 | 1.828 |
| France | after measures | 0.161 | 4.311 |
| Mexico | initial | 0.209 | 3.324 |

(\*) Not determinable

The last point is extremely important, since two drastically different slopes can be observed before and after a package of adequate measures within the same territory. In addition, two localities that experienced similar initial specific epidemic rates may exhibit dramatically different evolutions as a function of the initial response of government and society (Figure 1C,D). For instance, while the COVID-19 epidemics in Italy and South Korea exhibited practically equal µ_o_ values, the Italian outbreak has maintained the same growth rate throughout 20 days, while South Korea has set an example by effectively and rapidly lowering the specific epidemic rate to nearly 0 in just two weeks.

### Validation and predictions

We have run different scenarios to validate the predictive capabilities of our epidemic model for COVID-19. Overall, the model is capable of closely reproducing the progression of reported cases for urban areas of more than 5 × 10^6^ inhabitants (i.e., Iran, the city of Tehran in Iran, Spain, and NYC). We found that, adapting the model to a particular locality is straightforward and only requires (a) the calculation of the population and the surface area of the urban area, and (b) the selection of a t_d_ value (time to doubling the name of infections). Note that our model is formulated in terms of values of the specific epidemic growth rate (µ_o_ for the onset of the epidemic and µ for later times). However, expressing the specific epidemic rate in terms of doubling time (t_d_=Ln 2/µ) is more practical and simpler to communicate and understand (Table 2).

The selection of µ_o_ (t_d_) can be easily done by fitting the prediction to the initial set of reported cases of infection. In our experience, four to five reliable data points are needed for a good fit. For instance, Figure 2 shows the predicted trend of the pandemic in NYC during the first 28 days of March, 2020. In addition, we set (Dd/Dd_ref_=1.90), since the population density in NYC is 1.90-fold higher than that in Madrid. A value of t_d_= 2.25 was also set for the first week of this simulation. Later, at day 7 (March 7), we reset the value of t_d_ to 3.75 to reflect the modification of the slope of the local epidemic event in NYC (Figure 1d), due to the implemented measures of containment. Based on this exercise, we foresee that this simple modeling tool can be used to evaluate the efficacy of containment strategies. In other words, the value of µ_o_ required in the simulation to adapt the predicted data to the actual trend of the local epidemic provides an indicator of the local rate of spreading of the pandemic.

**Figure 2.**
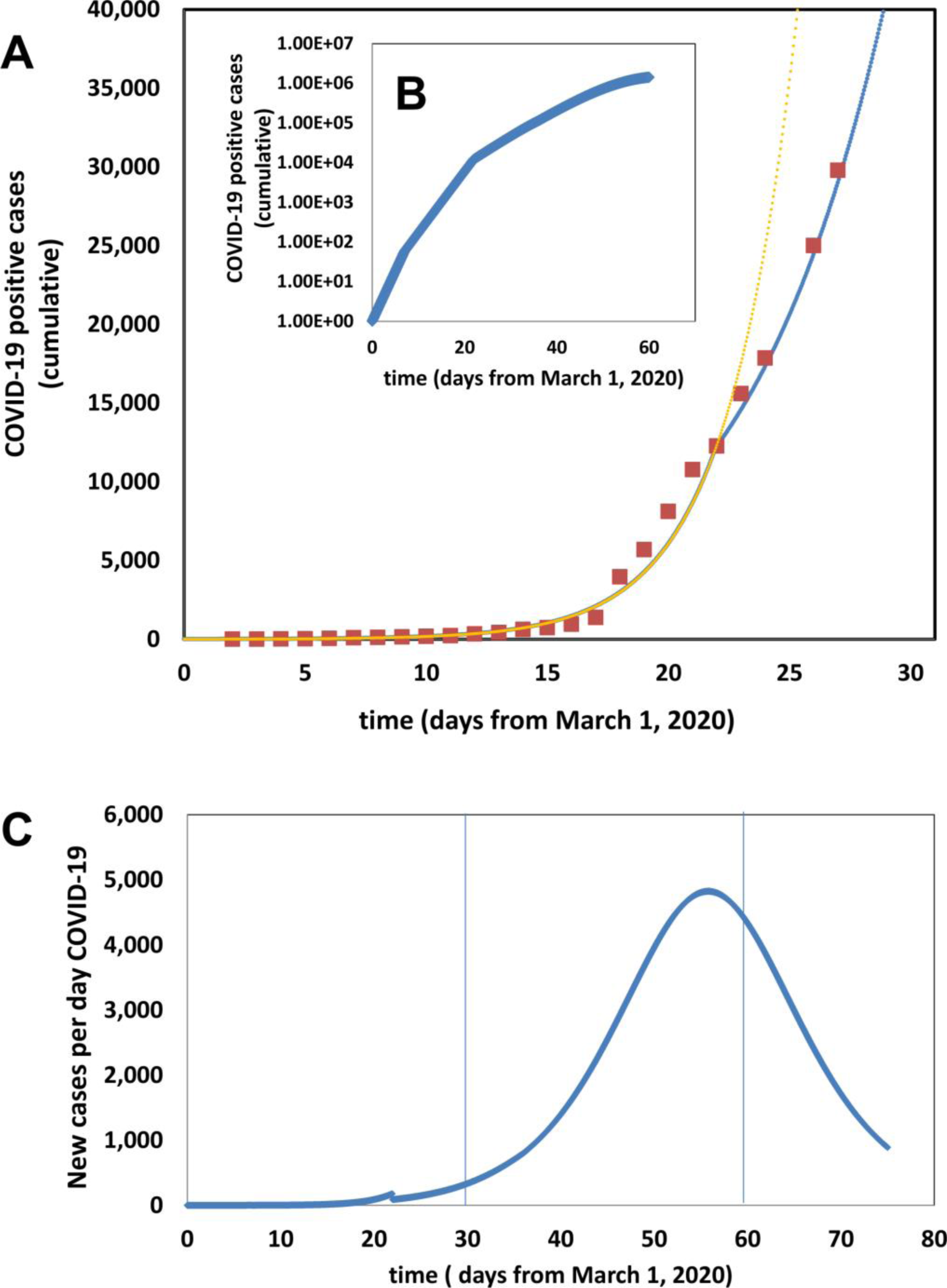
Progression of the COVID-19 Pandemic in NYC. (A) Initial evolution of the number of positive cases of COVID-19 in NYC. Actual data points, as officially reported, are shown using red circles. Simulation predictions are described by the blue dotted line. (B) Model prediction of the total number of symptomatic patients through the months of March and April. (C) Model prediction of new cases of COVID-19 during the period from March 1 to May 20, 2020 if no further containment actions are adopted.

Therefore, the differences between µ_o_ before and after interventions provide a real-time quantitative measure of the effectiveness of that set of measures. This can be extremely useful when assessing the efficacy of control of epidemics. For example, for NYC, this simple model states that the set of containment measures adopted during the first week of March in NYC diminished the specific rate of the epidemic by increasing the doubling time of infections from a value of 2.25 to 3.75 days.

The ability to make close predictions of the progression of cases in a particular region has profound and enabling implications. For example, in March 15^th^, our simulations predicted that, in absence of more aggressive containment measures (yellow trend in Figure 2A), the peak of infections in NYC will be reached by April 10, 2010, after reaching the unprecedented value of 11,000 new cases per day, and a cumulative number of 1 × 10^6^ citizens infected. However, we observed a deviation from this prediction by the third week of March that we attribute to the stringent measures of social distancing established in NYC earlier that week. Accordingly, we multiplied the value of (Dd/Dd_ref_) in our simulations by a factor of 0.50 to properly fit the new trend on actual cases (blue trend in Figure 2A). Note that his suggest that the measures of social distancing imposed in NYC were equivalent to decrease the effective demographic density to 50%. At the end of March, after this adjustment, our model forecasts a peak of infections of nearly 5,000 new cases per day (less than half than the prediction before social distancing), and a cumulative number of 1 × 10^6^ citizens infected.

### Effect of social distancing

Social distancing has been regarded as the one of the most effective buffering measures for local COVID-19 epidemics^30,31^. Next, we evaluate the effect of different degrees of social distancing on the shape of the epidemic curve for NYC, one of the most densely urban areas worldwide. This evaluation is straightforward, since the formulation of our model explicitly considers the demographic density of the region as the most important modifier of the rate of progression of the epidemics.

In the Excel implementation of the model, we multiply the demographic ration (Dd/Dd_ref_) by 0.75 to calculate the impact of distancing measures that would diminish social contact by 25%. Similarly, we multiply (Dd/Dd_ref_) by 0.50 to simulate the effect of a scenario of social distancing that would diminish close social interaction by 50%. Figure 4 shows the effect of three different degrees/levels of social distancing on the cumulative number of infections (Figure 3A) and on the number of new cases of infection per day (Figure 3B).

**Figure 3.**
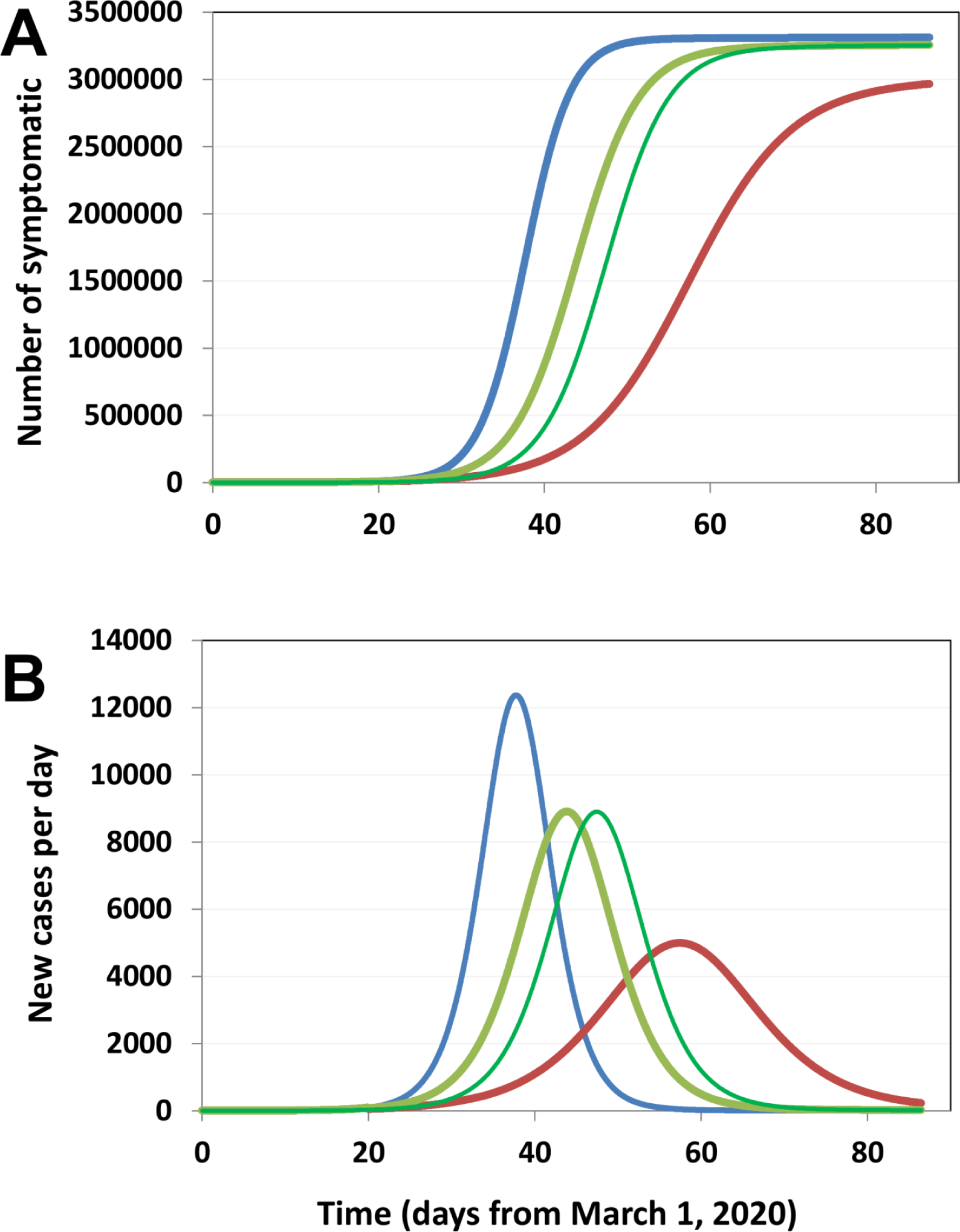
Prediction of the effect of social distancing on the progression of the COVID-19 pandemics in New York City (NYC). (A) Model prediction of the total number of symptomatic patients from March 1 to May 31, 2020 for different scenarios of social distancing: social distancing as in March 20, 2010 (blue line, current prediction); social distancing effective on March 20, whereby the effective demographic density in NYC is reduced by 25% (light green line); social distancing effective on March 20 whereby the effective demographic density in NYC is reduced by 50% (red line); and social distancing effective on March 10, whereby the effective demographic density in NYC is reduced by 25% (dark green line). (B) Model prediction for the number of new infections per day for each of the scenarios of social distancing described.

**Figure 4.**
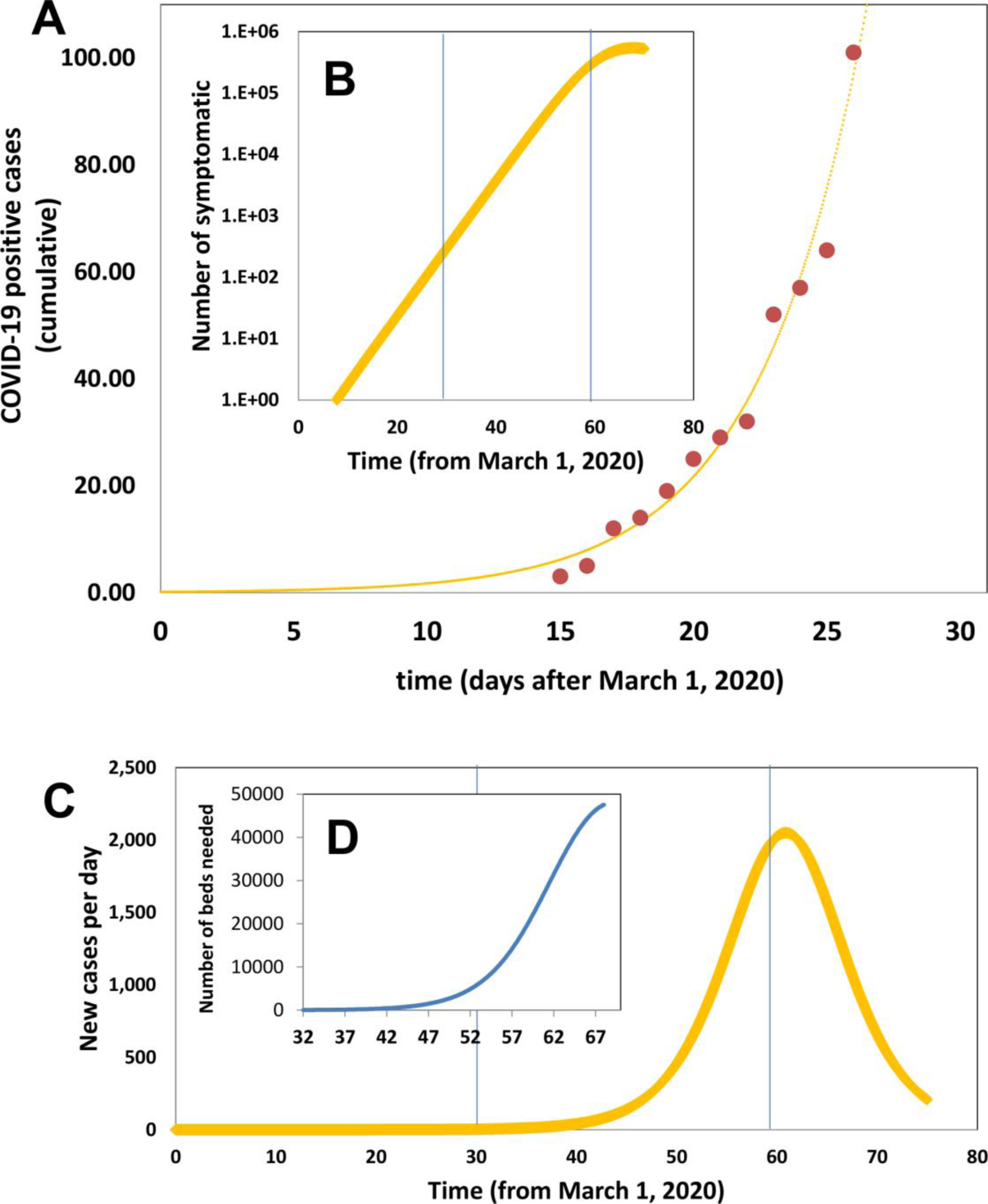
Progression of the COVID-19 pandemic in the metropolitan area of Monterrey, Nuevo León, Mexico. (A) Initial evolution of the number of positive cases of COVID-19 in the metropolitan area of Monterrey. Actual data points, as officially reported, are shown using red circles. Simulation predictions are described by the blue dotted line. (B) Model prediction of the total number of symptomatic patients from March 1 to May 20, 2020. (C) Model prediction of new cases of COVID-19 during the period from March 1 to May 20, 2020 if no further containment actions are adopted. (D) Estimation of the number of beds needed during the month of April 2020 in Monterrey, based on the number of patients that will require hospitalization according to the model predictions.

Social distancing has a clear buffering effect on the epidemics, delaying the occurrence of the peak of infections and distributing the number of cases across a longer time span. This is remarkably important as it provides time for proper attention to patients with severe symptomatology^5^.

For instance, our results suggest that, for an urban area such as NYC, imposing measures that guarantee a social distance equivalent to a decrease in demographic density of 50% will delay the peak of maximum number of infections by 25 days and will decrease its intensity from 23000 to 9000 new cases of infection per day. In turn, this implies a lower demand for hospital beds per day during the epidemics and may mark the difference between a manageable crisis and a public health catastrophe^5,30^.

Interestingly, the effect of anticipating measures of social distancing has a moderate effect on retarding the infection curve but not on decreasing the cumulative number of infections. This moderate gain of time provides additional leeway for planning interventions or allocating resources, with time being gold during pandemic events.

### Prediction in real time

We are currently following the onset of the COVID-19 pandemic in Monterrey, the second most industrialized city in México and the third most populated. Monterrey has a similar demographic density to that of Madrid (Dd/Dd_ref_=0.95). In addition, we set t_d_ = 2.5, based on proper fitting to the first set of official values of COVID-19 infected announced for Monterrey by the local authorities from March 15 to March 19, 2020. Remarkably, the simulation results have accurately predicted the nine subsequent actual values, as officially reported from March 19 to March 28 (Figure 4 A). Monitoring actual data, while comparing with model predictions, enables real time assessment of the effectiveness of the containment measures. In turn, this empowers officials, scientists, health care providers, and citizens. Moreover, friendly and widely available mathematical modeling enables rational planning. For instance, according to the pandemic scenario predicted for Monterrey, in the absence of further containment measures and stricter social distancing, the total number of symptomatic infected will surpass 650,000 persons (Figure 4B), and the number of new infections per day (Figure 4C) will exhibit a peak of 2000 by the end of April. The simulation may be used to forecast the demand of beds during the month of April 2020, which is estimated to exhibit a peak of nearly 50000.

This estimate considers that only 10.0% of the symptomatic patients will require hospitalization, which may be optimistic. Reports based in 44000 COVID-19 cases in China indicate that the percentage of patients with severe symptoms may be 14%, with a 5% of critical cases^32^. In prospective, the total number of beds in the Mexican public health sector is estimated in 20,000 (for the whole country).

### Concluding remarks

We used a set of differential equations, recent epidemiological data regarding the evolution of COVID-19 infection in a reduced set of regions (i.e., Spain, Iran, and NYC), and basic information on the characteristics of COVID-19 infection (i.e., time from infection to recovery, case mortality rate) to accurately recreate the onset of the COVID-19 in two urban areas with different demographic characteristics (i.e., NYC and Monterrey, México). We showed that the model can be adapted to closely follow the evolution of COVID-19 in densely populated urban areas by simply adjusting two parameters related to (a) population density and (b) aggressiveness of the response from a society/government to epidemics.

Scenarios such as those currently unfolding in Iran, Italy, or Spain emphasize the importance of planning ahead during epidemic events. The availability of a simple model may be highly enabling for local governments, physicians, civil organizations, and citizens as they struggle in their endeavor to accurately forecast the progression of an epidemic and formulate a plan of action. As previously stated, the use of simple/user-friendly models to evaluate in (practically) real time the effectiveness of containment strategies or programs may be a powerful tool for analyzing and facing epidemic events^6,12^. This contribution shows the prediction potential of an extremely simple simulation tool that can be used by practically any citizen with basic training in Excel.

## Data Availability

All data required to (a) validate the information presented in the manuscript and use the model presented in the manuscript is available. Relevant links are included in the text. The Excel spreadsheet required to run the model is available as supplementary material.

https://elpais.com/sociedad/2020/03/16/actualidad/1584360628_538486.html

## Acknowledgments

MMA, EGG, and GTdS acknowledge the funding received from CONACyT (Consejo Nacional de Ciencia y Tecnología, México).

## Author contributions

MMA, EGG, and GTdS collected and analyzed epidemiology data. MMA formulated the model and run the simulations. MMA and GTdS wrote the manuscript. All authors reviewed and approved the manuscript.

## References

1. Holshue, M. L. et al. First Case of 2019 Novel Coronavirus in the United States. N. Engl. J. Med. (2020). doi:10.1056/nejmoa2001191

2. MacIntyre, C. R. Global spread of COVID-19 and pandemic potential. Glob. Biosecurity 1, (2020).

3. Choi, S. C. & Ki, M. Estimating the reproductive number and the outbreak size of Novel Coronavirus disease (COVID-19) using mathematical model in Republic of Korea. Epidemiol. Health e2020011 (2020). doi:10.4178/epih.e2020011

4. Wong, J. E. L., Leo, Y. S. & Tan, C. C. COVID-19 in Singapore-Current Experience: Critical Global Issues That Require Attention and Action. JAMA (2020). doi:10.1001/jama.2020.2467

5. Remuzzi, A. & Remuzzi, G. COVID-19 and Italy: what next? Lancet (2020). doi:10.1016/s0140-6736(20)30627-9

6. Roosa, K. et al. Real-time forecasts of the COVID-19 epidemic in China from February 5th to February 24th, 2020. Infect. Dis. Model. 5, 256–263 (2020).

7. Peng, L., Yang, W., Zhang, D., Zhuge, C. & Hong, L. Epidemic analysis of COVID-19 in China by dynamical modeling. (2020).

8. Kucharski, A. J. et al. Early dynamics of transmission and control of COVID-19: a mathematical modelling study. Lancet Infect. Dis. (2020). doi:10.1016/S1473-3099(20)30144-4

9. Jung, S. et al. Real-Time Estimation of the Risk of Death from Novel Coronavirus (COVID-19) Infection: Inference Using Exported Cases. J. Clin. Med. 9, 523 (2020).

10. Hellewell, J. et al. Feasibility of controlling COVID-19 outbreaks by isolation of cases and contacts. Lancet Glob. Heal. 8, e488–e496 (2020).

11. Gostic, K., Gomez, A. C. R., Mummah, R. O., Kucharski, A. J. & Lloyd-Smith, J. O. Estimated effectiveness of symptom and risk screening to prevent the spread of COVID-19. Elife 9, (2020).

12. Cauchemez, S., Hoze, N., Cousien, A., Nikolay, B. & ten bosch, Q. How Modelling Can Enhance the Analysis of Imperfect Epidemic Data. Trends in Parasitology 35, 369–379 (2019).

13. Bao, L. et al. Reinfection could not occur in SARS-CoV-2 infected rhesus macaques. bioRxiv 2020.03.13.990226 (2020). doi:10.1101/2020.03.13.990226

14. Prompetchara, E., Ketloy, C. & Palaga, T. Allergy and Immunology Immune responses in COVID-19 and potential vaccines: Lessons learned from SARS and MERS epidemic. doi:10.12932/AP-200220-0772

15. Liu, W. et al. Two-Year Prospective Study of the Humoral Immune Response of Patients with Severe Acute Respiratory Syndrome. J. Infect. Dis. 193, 792–795 (2006).

16. Zhou, P. et al. A pneumonia outbreak associated with a new coronavirus of probable bat origin. Nature 579, 270–273 (2020).

17. Mizumoto, K., Kagaya, K., Zarebski, A. & Chowell, G. Estimating the asymptomatic proportion of coronavirus disease 2019 (COVID-19) cases on board the Diamond Princess cruise ship, Yokohama, Japan, 2020. Eurosurveillance 25, 2000180 (2020).

18. Nishiura, H. et al. Estimation of the asymptomatic ratio of novel coronavirus infections (COVID-19). medRxiv 2020.02.03.20020248 (2020). doi:10.1101/2020.02.03.20020248

19. Leung, N. H. L., Xu, C., Ip, D. K. M. & Cowling, B. J. The fraction of influenza virus infections that are asymptomatic: a systematic review and meta-analysis. doi:10.1097/EDE.0000000000000340

20. Elizondo-Montemayor, L. et al. Seroprevalence of antibodies to influenza A/H1N1/2009 among transmission risk groups after the second wave in Mexico, by a virus-free ELISA method. Int. J. Infect. Dis. 15, e781–e786 (2011).

21. Zimmer, S. M. et al. Seroprevalence Following the Second Wave of Pandemic 2009 H1N1 Influenza in Pittsburgh, PA, USA. doi:10.1371/journal.pone.0011601

22. Lan, L. et al. Positive RT-PCR Test Results in Patients Recovered From COVID-19. JAMA (2020). doi:10.1001/jama.2020.2783

23. Bi, Q. et al. Epidemiology and Transmission of COVID-19 in Shenzhen China: Analysis of 391 cases and 1,286 of their close contacts. medRxiv 2020.03.03.20028423 (2020). doi:10.1101/2020.03.03.20028423

24. Zou, L. et al. SARS-CoV-2 Viral Load in Upper Respiratory Specimens of Infected Patients. N. Engl. J. Med. 382, 1177–1179 (2020).

25. Bai, Y. et al. Presumed Asymptomatic Carrier Transmission of COVID-19. JAMA (2020). doi:10.1001/jama.2020.2565

26. Lai, C. C., Shih, T. P., Ko, W. C., Tang, H. J. & Hsueh, P. R. Severe acute respiratory syndrome coronavirus 2 (SARS-CoV-2) and coronavirus disease-2019 (COVID-19): The epidemic and the challenges. International Journal of Antimicrobial Agents 55, 105924 (2020).

27. Xu, Z. et al. Pathological findings of COVID-19 associated with acute respiratory distress syndrome. Lancet Respir. Med. 0, (2020).

28. Porcheddu, R., Serra, C., Kelvin, D., Kelvin, N. & Rubino, S. Similarity in Case Fatality Rates (CFR) of COVID-19/SARS-COV-2 in Italy and China. J. Infect. Dev. Ctries. 14, 125–128 (2020).

29. Linton, N. M. et al. Epidemiological characteristics of novel coronavirus infection: A statistical analysis of publicly available case data. doi:10.1101/2020.01.26.20018754

30. Anderson, R. M., Heesterbeek, H., Klinkenberg, D. & Hollingsworth, T. D. How will country-based mitigation measures influence the course of the COVID-19 epidemic? The Lancet 395, 931–934 (2020).

31. Isolation, quarantine, social distancing and community containment: pivotal role for old-style public health measures in the novel coronavirus (2019-nCoV) outbreak | Journal of Travel Medicine | Oxford Academic. Available at: https://academic.oup.com/jtm/article/27/2/taaa020/5735321. (Accessed: 24th March 2020)

32. Wu, Z. & McGoogan, J. M. Characteristics of and Important Lessons from the Coronavirus Disease 2019 (COVID-19) Outbreak in China: Summary of a Report of 72314 Cases from the Chinese Center for Disease Control and Prevention. JAMA - J. Am. Med. Assoc. (2020). doi:10.1001/jama.2020.2648

